# Relating mutational signature exposures to clinical data in cancers via signeR 2.0

**DOI:** 10.1101/2023.04.12.23288466

**Authors:** Rodrigo Drummond, Alexandre Defelicibus, Mathilde Meyenberg, Renan Valieris, Emmanuel Dias-Neto, Rafael A. Rosales, Israel Tojal da Silva

## Abstract

**Motivation:** Cancer is a collection of diseases caused by the deregulation of cell processes, which is triggered by somatic mutations. The search for patterns in somatic mutations, known as mutational signatures, is a growing field of study that has already became a useful tool in oncology. Several algorithms have been proposed to perform one or both the following two tasks: 1) *de novo* estimation of signatures and their exposures, 2) estimation of the exposures of each one of a set of pre-defined signatures. Our group developed signeR, a Bayesian approach to both these tasks.

**Results:** Here we present a new version of the software, signeR 2.0, which extends the possibilities of previous analyses to explore the relation of signature exposures to other data of clinical relevance. signeR 2.0 includes an user-friendly interface developed using the R-Shiny framework and improvements in performance. This version allows the analysis of submitted data or public TCGA data, which is embedded on the package for easy access.

**Availability:** signer 2.0 is an open-source R package available through the Bioconductor project at https://doi.org/doi:10.18129/B9.bioc.signeR

## Introduction

DNA mutations accumulate throughout an individual’s life and may result in the deregulation of metabolic processes observed in tumor cells (Stratton, 2011). Specific patterns of somatic mutations are characteristic of the exposure to some carcinogens, which are more frequently found in some tumor types. The study of these ‘mutational signatures’ has become a solid field of research in oncology, and is now seen as a field which has made significant advances over the last years (Alexandrov, Nik-Zainal, Wedge, et al., 2013; Koh et al., 2021). The importance of studying mutational signatures in oncology is irrefragable, as mutation patterns are related to cancer aetiology, diagnosis and prognosis, appear to predict response to therapy (Liu, Xia, et al., 2022; Liu, Lin, et al., 2022) and may echo genomic alterations induced by chemotherapy, making the valuable tools for most aspects of cancer research Koh et al., 2021.

The first method to extract mutational signatures from somatic mutation counts was based on non-negative matrix factorisation (NMF) techniques applied to Single Nucleotide Variations (SNVs) counts, see Alexandrov, Nik-Zainal, Wedge, et al., 2013. Since then, several methods for mutational signature extraction have emerged, most of them based on variations of the NMF algorithm; see Kim et al., 2021 for a recent overview and a comparison of current methods. Our group developed signeR, a Bayesian approach to the NMF paradigm for mutational signature extraction, Rosales et al., 2017. A key idea that led to the development of signeR is that the signature extraction problem can be treated as an inferential task, subject to statistical modelling. signeR is able to extract the underlying signatures by estimating both the number of signatures present in the data and the relative contribution of each signature to the total amount of observed mutation counts. The relative contribution of a signature to the total amount of counts is known as a signature exposure. signeR can also be used to estimate the sample exposure levels of known mutational signatures, such as those described by the COSMIC consortium (Tate et al., 2018) or the Signal initiative (Degasperi et al., 2020). This functionality follows a tendency observed in literature: as signatures have become known and well determined by the study of extensive datasets, algorithms capable of fitting mutation samples to available signatures started to emerge (*e*.*g*. deconstructSig, Rosenthal et al., 2016).

Mutational signatures have recently been proposed as markers for cancer prognosis or drug sensitivity (see reviews by Brady, Gout, and Zhang, 2022 and Levatic et al., 2022). Available evidence suggests that the estimation of exposure levels to mutational processes may be incorporated within the cancer diagnostic workflow, which may improve diagnosis in the future Van Hoeck et al., 2019. As an example, our group recently considered signeR to stratify gastric cancer patients for therapeutic intervention (Buttura et al., 2021). Those results highlight the scientific potential of relating mutational signatures to other relevant features in cancer, such as clinical or molecular data.

In this article we describe an enhanced version of signeR that is computationally more efficient and has several new functionalities. A major contribution of signeR 2.0 is that it allows to study the relation of each signature exposure to almost any other clinical feature of interest, such as overall survival, tumor staging or cancer subtypes. These features may be categorical (*e*.*g*. cancer molecular subtypes), continuous (*e*.*g*. gene expression) or survival data. Such additional information is nowadays present in several data bases as for instance in The Cancer Genome Atlas consortium (TCGA, Zhang et al., 2021). Clustering or machine learning algorithms used to relate exposures to clinical features are repeatedly applied to different results obtained while estimating the matrix of exposures to signatures. The decomposition of mutation data may lead to multiple similarly suitable solutions, thus the estimation of signatures and exposures is not exact. Most publications use bootstrap methods to evaluate the robustness of results obtained from mutation data decomposition (Alexandrov, Nik-Zainal, Siu, et al., 2015; Huang, Wojtowicz, and Przytycka, 2018). Our method, however, employs a Gibbs sampler to generate a posterior distribution of estimate signatures and exposures.

The utility of signeR 2.0 is demonstrated here by considering TCGA data obtained from stomach adenocarcinomas. Mutational signatures previously identified by the COSMIC consortium (Tate et al., 2018) for this type of cancer were used as templates to correlate their observed exposures to several other clinical data of interest. These analyses include the clustering of samples according to signatures exposures, the search for signatures showing significant differences in distribution among tumor subtypes and the evaluation of how exposure levels affect patients overall survival.

The software interface is user-friendly and intuitive, facilitating the estimation of mutational signatures and further extending the study of their relation to other clinical data to users with little programming background. We hope that this version of signeR will aid in subsequent genome based studies of cancers, eventually leading to new insights and discoveries.

## System and methods

### Database content

The new version of signeR described here provides a query interface, signerRFlow, to explore the interplay of mutational signature exposures and several other features present in clinical data. To do so, signeR 2.0 embedded into its framework the most recently processed and up-to-date molecular and clinical dataset of The Cancer Genome Atlas (TCGA) consortium (https://www.cancer.gov/about-nci/organization/ccg/research/structural-genomics/tcga) along with a catalog of mutational signatures (COSMIC Single Base Substitution signatures v3.2, the latest version by the software construction, https://cancer.sanger.ac.uk/cosmic/signatures/SBS).

### Interface design

The signeRFlow app was developed using shiny Chang et al., 2022, an R package for building interactive web apps. It is implemented as an open-source R package available along with signeR 2.0 through the Bioconductor project at https://doi.org/doi:10.18129/B9.bioc.signeR.

### Algorithm

signeR 2.0 presents an updated version of the signeR Bayesian approach Rosales et al., 2017, with parallel computation capabilities devoted to hasten processing time. The hyper-hyper parameters of our Bayesian hierarchical model have been estimated for the TCGA data. This saves further computational time and resources. Nevertheless, as in previous versions, there is still an option to estimate the hype-hyper parameters while inferring signatures and their exposures.

To further explore the genotype-phenotype relationships between mutational signatures and other data of interest, signeR 2.0 provides an unified data modeling toolkit. If additional samples information is available, including molecular and clinical data such as cancer sub-type or overall survival, signeR 2.0 is able to evaluate how this information relates to the estimated exposures to mutational signatures. When the additional data is of a categorical nature, differences in exposures among groups can be analyzed and, if some of the samples are unlabeled they can be labeled based on the similarity of their exposure profiles to those of labeled samples. In the case of a continuous additional feature, its correlation to estimated exposures can be evaluated. Survival data can also be analyzed by estimating the relation of survival to mutational signature exposure. We describe briefly each of these new features next.

signeR takes as input a matrix ***M*** = (***M***_*ij*_) of mutation counts found in a set of genome samples. Each column of ***M***, denoted hereafter as ***M***_*j*_, corresponds to a genome sample and each row to a given mutation type. As an output signeR can estimate two matrices ***P*** and ***E*** of mutation signatures and signature exposures such that ***M*** ≈ ***PE***. Alternatively, signeR can estimate only the exposures ***E*** to known signatures. In both cases, the algorithm estimates exposures by drawing a sample ***E***^(1)^, ***E***^(2)^, …, ***E***^(*R*)^ of exposure matrices, approximately distributed according to our model posterior distribution (Rosales et al., 2017). All subsequent analyses described here are based on the repeated application of statistical or learning algorithms to the matrices ***E***^(*r*)^, 1 ≤ *r* ≤ *R*. After each of the sampled matrices ***E***^(*r*)^ is analyzed, results are joined and findings are considered significant if they are consistent throughout most of these analyses. A general description of this procedure is shown by the pseudo-code presented in Algorithm 1.

If estimated exposures are confronted to a categorical feature, signeR 2.0 uses non-parametric tests (Wilcoxon-Mann-Whitney or Kruskal-Wallis tests) to assess the enrichment of exposures in any of the categories. For each signature, the tests are applied on each ***E***^(*r*)^, and obtained *p*-values are inverted and log-transformed for visualisation purposes. Resulting values are called Differential Exposure Scores and can be visualized as a boxplot (for more details see Rosales et al., 2017). signeR 2.0 is also able to evaluate the ability of exposure levels to discriminate samples among categories. Several classification algorithms are available for this purpose. signeR currently includes: 1. *k*-nearest neighbors, 2. linear vector quantization, 3. Logistic regression, 4. linear discriminant analysis, 5. least absolute shrinkage and selection operator (lasso), 6. naive Bayes, 7. support vector machines, and 8. random forests. In all cases, given a genome sample ***M***_*j*_ and a exposure matrix ***E***^(*r*)^ the chosen classifier is used to label ***M***_*j*_ in one of the categories. The final label for ***M***_*j*_ is obtained as the most frequent label obtained by considering ***E***^(*r*)^, 1 ≤ *i* ≤ *R*.

#### Algorithm 1 Exposure data analysis: general structure

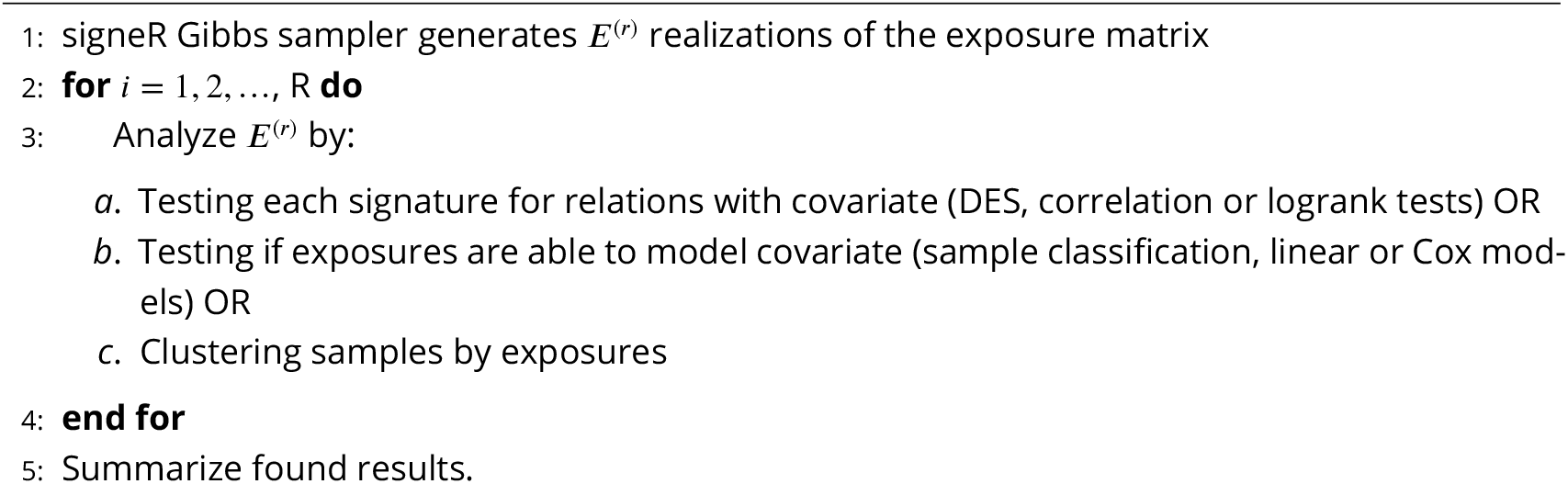

When a continuous feature is considered, such as gene expression, the correlation of each signatures exposure to this feature can be assessed. A correlation test is applied to each ***E***^(*r*)^ and the found *p*-values, inverted and log-transformed, are shown as a boxplot. A similar approach, considering all signatures together, is used by signeR 2.0 to evaluate whether the feature can be linearly modeled based on exposures.

Survival data, often present in cancer studies, can also be related to exposures. For each signature and each ***E***^(*r*)^, signeR 2.0 stratifies patients according to exposure levels and applies logrank tests to compare obtained groups. The impact of exposures on survival can also be quantified via Cox proportional hazard models (Therneau and Grambsch, 2000). Again, all tests are applied to each ***E***^(*r*)^ and results are summarized by taking the median of all the obtained statistics (*p*-values and hazard ratios).

Finally, when no additional data is available, signeR 2.0 includes unsupervised methods such as hierarchical and fuzzy clustering to discover sample sub-groups based entirely on the estimated exposures. Several options are available for the required distance measure (see R function dist documentation) or the agglomerative procedure (see R function hclust documentation). If a hierarchical clustering is used, the algorithm is applied to each exposure matrix ***E***^(*r*)^, 1 ≤ *i* ≤ *R*, as mentioned in the pseudo-code Algorithm 1. The obtained dendrograms are compared and shown on a final chart, were the relative frequency with which each branch were found is displayed. In case the user chooses to use fuzzy clustering, the fuzzy C-means algorithm is applied to each ***E***^(*r*)^, thus generating matrices of membership grades of each genome sample to each cluster. Those grades are averaged to yield the final result. For visualisation purposes a hierarchical procedure is applied to the mean membership grades so that similar samples are displayed together on the final chart.

Tests and learning algorithms available on signeR 2.0 are obtained from specialized R packages (e.g. pvclust or survival). Their complete list can be found on the package documentation and is included as Supplementary Material. Few examples of the application of these functionalities to a data set from TCGA data base are presented in Section.

### Implementation: signeRFlow

The signeRFlow app includes three major components and consists of a pipeline that allows: (i) data input and pre-processing; (ii) mutational signature estimation or fitting and (iii) exposure data modeling. A schematic overview of signerFlow is shown in Figure 1.

**Figure 1.**
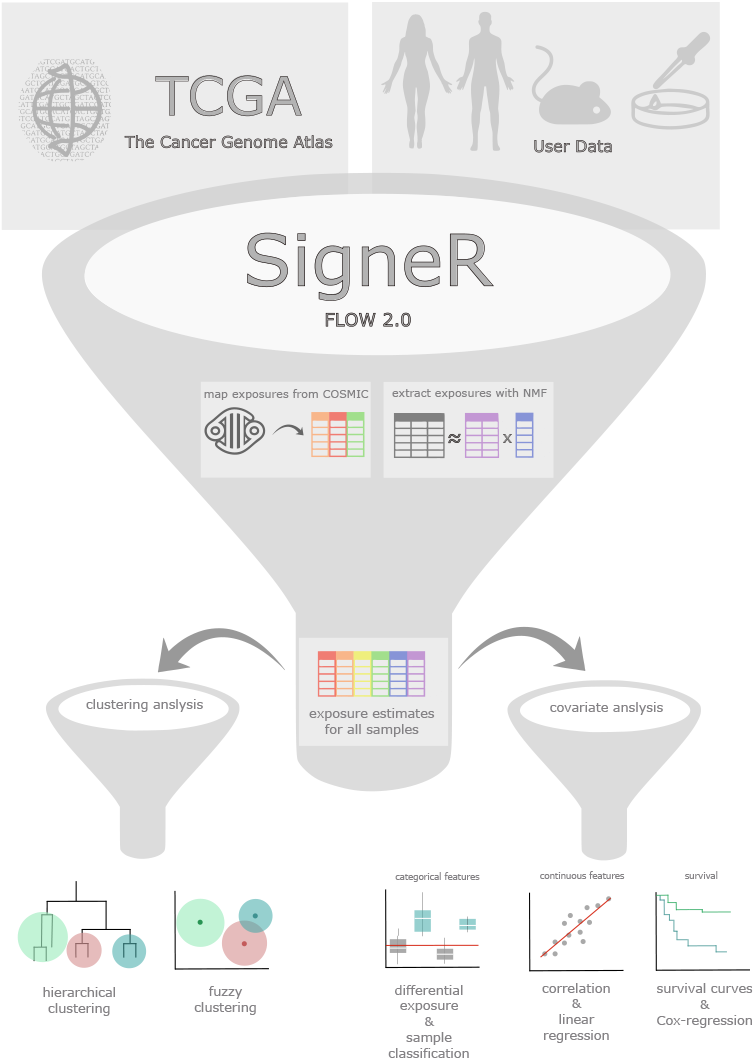
General overview of SigneRFlow. Starting from the top, clinical and molecular features from publicly available TCGA databases or own user data can be loaded. After pre-processing, a friendly interface provides options to setup both *de novo* and *fitting* analyses. After pre-processing, a friendly interface provides analytical methods for downstream analysis.

The flexible input interface was designed to allow users to upload their own data either as VCF file or a SNV matrix file (Smf, an example of the file structure can be found within the interface). Additionally, the user can select a previous cancer study of interest from the TCGA database available in the *TCGA Explorer* module. In the first option, users can add clinical information, while available clinical data for TCGA samples is already organized within signeRFlow and easily accessible through the interface.

Upon data upload completion, the mutational signature analysis is ready to commence. In this step, the user can take advantage of a Bayesian approach to perform *de novo* identification of mutational signatures. signeR 2.0 provides flexible options for choosing the number of searched signatures or optimizing it, within a fixed range, according to the Bayesian Information Criterion (BIC). In addition, signeRFlow is able to fit the mutational spectra of studied genome samples to known mutational signatures, thus estimating the samples exposure levels to related mutational processes. Single Base Substitution (SBS) signatures from COSMIC are available within signeRFlow for *fitting* analysis, although users can upload other signatures as well. Whenever a mutational signature analysis is performed, signeRFlow offers several plot options to visualize estimated signatures and their exposures, as well as the convergence of the MCMC model used to estimate them (Supplementary Figure 5). For the fitting to known signatures, exposure plots are available (see, for instance, Figure 2A).

**Figure 2.**
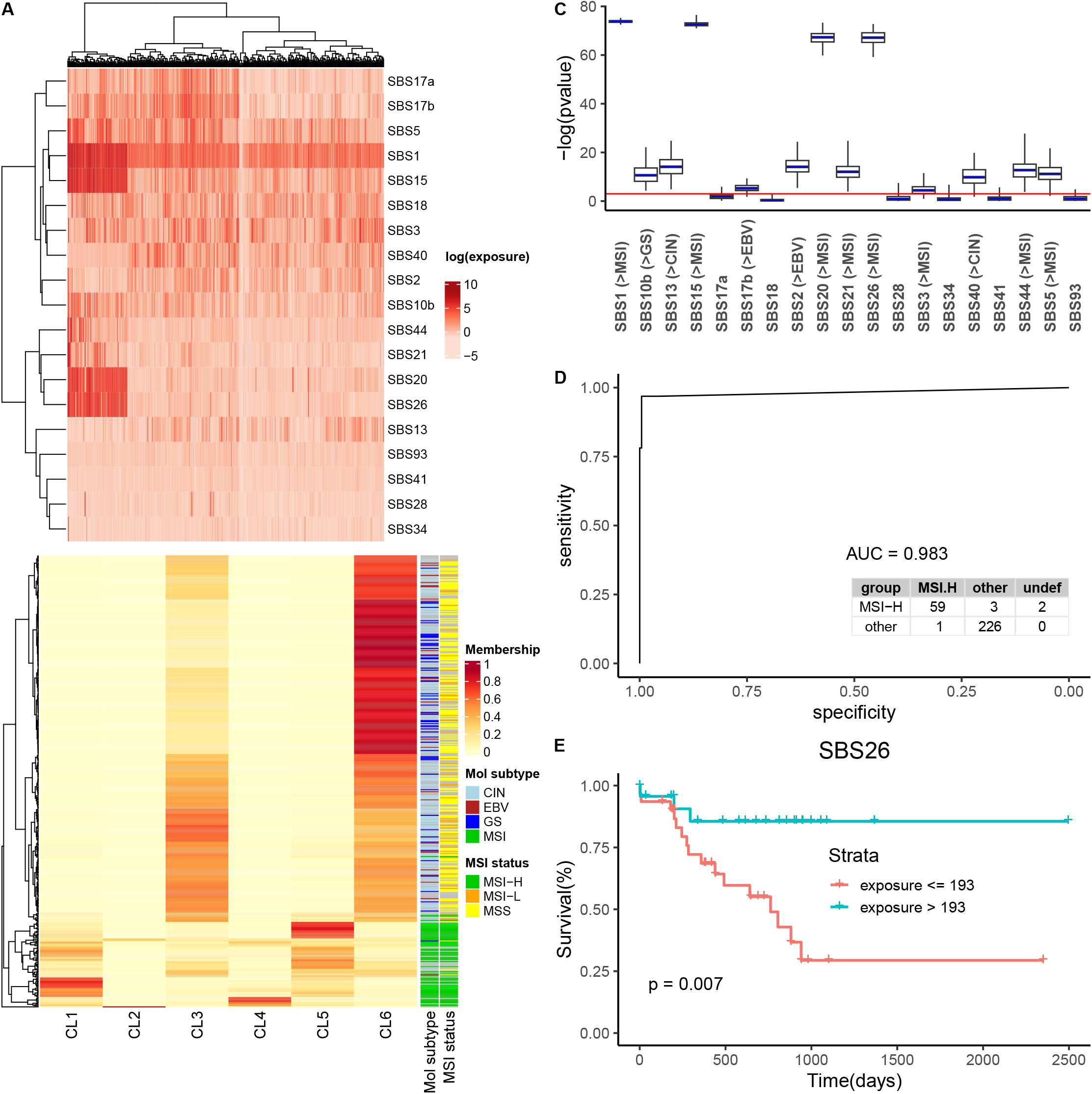
A) Heatmap of estimated exposures obtained by *fitting* 19 COSMIC signatures to the STAD dataset. Genome samples are displayed as columns of the heatmap while COSMIC signatures are arranged as rows and estimated (log-transformed) exposures levels are shown by the colour scale. B) Fuzzy clustering of samples according to estimated exposures, compared to known classifications by molecular profiles. Clusters were organized in columns and for each sample (row) the colour code indicates the membership grade to each cluster. Following the fuzzy clustering approach, a hierarchical clustering algorithm was applied to the membership grades (dendrogram at left), enabling better visualisation of results and allowing to establish a relation to molecular sub-types and MSI status (annotation columns at right side). C) *p*-values found by Kruskal-Wallis test for differences in exposures among the four sample groups. For comparison and display purposes, the *p*-values were inverted and log-transformed. Box-plots of obtained scores are displayed and the signficance cutoff of 0.05 is indicated by the red line. The labels at the x axis correspond to the id of each signature and, for those showing significant differences, the group characterized by higher exposure levels. D) ROC curve of the exposure-based classification of samples according to their MSI status and related confusion matrix. E) Kaplan-Meier curves showing the overall survival of STAD patients after stratification by the exposures obtained while fitting COSMIC signature SBS26. The displayed *p*-value was found by application of the log-rank test for defined sample groups.

Finally, signeRFlow provides a toolbox containing state of art techniques on learning algorithms for exposure data analysis (see Algorithm 1). For example, hierarchical and fuzzy clustering can be used to explore the qualitative differences among samples evidenced by signature exposures. Furthermore, to unveil the interplay of mutational signatures with clinical or genomic features, signeRFlow provides comprehensive options for covariate analysis considering either categorical or numerical features. In the first case, signeR 2.0 *Differential Exposure Score* (DES) can highlight signatures that are differentially active among previously defined groups of samples, while the function *ExposureClassify* evaluates the assignment of samples to groups according to exposure profiles. On the other hand, sample correlation and linear regression can be performed. Lastly, the effect of exposure levels on prognosis can be investigated by comparing the survival distributions of sample groups with contrasting exposure levels or by Cox regression analysis. The next section presents a concrete example of these possibilities.

### A case study

Although standard histological classification techniques are fundamental for dividing cancer into sub-types and disease stratification, the exposure to mutational processes may provide additional information extending further this characterization. We illustrate this here by using the differential exposure score (DES, Rosales et al., 2017) estimated while analysing a data set with 439 samples selected from the stomach adenocarcinoma (STAD) cohort from TCGA. The mutational spectra of these samples were fitted to known signatures previously reported to be characteristic of STAD (Alexandrov, Nik-Zainal, Siu, et al., 2015). According to COSMIC nomenclature, the signatures included in this analysis are **S**ingle **B**ase **S**ubstitutions (SBS) numbers 2, 3, 5, 10b, 13, 15, 17a, 17b, 18, 20, 21, 26, 28, 34, 40, 41, 44, and 93. The estimated exposures (i.e. the empirical average of the realizations obtained by signeR for the exposure matrix) are shown in Figure 2A.

As an exploratory approach, a Fuzzy clustering algorithm was applied to the exposures found by signeR. Results are shown in Figure 2B. Interestingly, 3 of the 6 groups found via fuzzy clustering (Figure 2B, clusters 1, 4 and 5) are mainly composed of samples characterized by high microsatellite instability (MSI), an important marker for tumour prognosis (Bass, 2014).

Motivated by the clustering results, we considered several supervised approaches available on signeR 2.0. The sample molecular sub-types proposed in Bass, 2014, namely Epstein-Barr virus (EBV)-positive tumours, tumours characterized by microsatellite instability (MSI), genomically stable (GS) tumours and tumours showing chromosomal instability (CIN), were adopted as targets to evaluate how the exposures of individual signatures correlate to them. For each signature, differences in exposures among STAD sample groups were evaluated by the Kruskal-Wallis test (*Differential Exposure Scores*). Results are shown in Figure 2C. Thirteen COSMIC signatures show different levels of activity in sample subtypes. Among signatures with higher exposures in MSI samples we found SBS1, a clock-like signature which in most cancers correlates with the age of the individual, and five mutational signatures associated with defective DNA mismatch repair and microsatellite instability: SBS15, SBS20, SBS21, SBS26 and SBS44 (COSMIC consortium).

The potential of exposure data to classify cancer samples was also tested in signeRFlow, based on the microsatellite instability (MSI) status also described by Bass, 2014. According to clustering and DES results, exposure data seems adequate to identify samples with high microsatellite instability. Thus, the original sample classification as **MSI-H**igh, **MSI-L**ow and **MSS**table was grouped as MSI-High and others and the classification algorithm adopted this grouping as target. A *k*-fold cross validation approach (*k*=8) was adopted, producing a ROC curve for the classification found, as well as the related confusion matrix (Figure 2D). It is worth noting that, as shown in the last column of the confusion matrix, a few samples are not consistently classified by signeR 2.0 and therefore are considered as *undefined*. Although the fraction of these samples is small (< 0.69%), their labeling to some group could be spurious, which is avoided by our approach because it incorporates the variability of exposure data.

Finally, we considered the impact of signature exposure levels on disease prognosis. For each signature, samples were stratified by their exposure levels, after searching for the cutoff value leading to the most relevant contrast on the overall survival of found strata (function maxstas, R package maxstat). The survival contrast among the resulting groups was evaluated by the logranktest, repeatedly applied to the realizations of the exposure matrix. Signatures SBSx, x = 1, 5, 15, 21 and 26 were reported as significant in prognosis. According to COSMIC, the first two are clock-like signatures, which correlates with the age of the individual, while the last three are associated with MSI samples. As an example, Kaplan-Meier survival curves for signature SBS26 can be found on Figure 2E.

The results presented in this section are consistent with previous knowledge about STAD. They exemplify how the new signeR functionalities described here can be used to gain further insights about the molecular nature of cancers.

## Discussion

signeR 2.0 is a software suite devoted to explore the information obtained from exposure to mutational processes data. It offers an updated version of the signeR Bayesian approach, with parallel computation functionalities and pre-computed hyper-hyper parameters, which saves computational time. It is presented in an user interface, signeRFlow, which brings in a ready-to-use form methods to estimate exposure data from mutation counts and to relate them with available clinical data from genome samples under study. The results of previous applications of signeR to the TCGA datasets, both *de novo* and fitting analyses, are available for exploration with signeR 2.0 tools, accompanied by related clinical data. To this end, signeR 2.0 offers a collection of established data analysis methods (classifiers, linear models, survival analysis, etc.) and interfaces to apply them to generated samples of the exposure matrix, outputting summary statistics of individual results.

Results found on the gastric adenocarcinoma dataset (TCGA-STAD) show the software’s potential for exploring available data, hopefully leading to further insights and new discoveries. The observed relation of exposures to some signatures and MSI status or age is in accordance with the literature (Bass, 2014) and demonstrates the potential of this tool to identify patterns of interest in cancer samples. Provided algorithms can be valuable tools to improve patient stratification or prognosis. Due to its software interface, signeRFlow, the use of signeR 2.0 does not require extensive computational training and therefore the tool is accessible for a wider audience. signer 2.0 is available as a Bioconductor package. A detailed explanation about how to use its interface is provided as Supplementary material (S1) and also in the package documentation. signeR is an ongoing project and new versions and functionalities will be released soon.

## Data Availability

All data produced are available online at https://doi.org/doi:10.18129/B9.bioc.signeR

