## Supplementary material for "Relating mutational signature exposures to clinical data in cancers via signeR 2.0"

### signeRFlow - Supplementary material

April 12, 2023

#### 1 Introduction

signeRFlow is a shiny app that allows users to explore mutational signatures and exposures to related mutational processes. With available modules, users are able to perform analysis on their own data applying different approaches, *de novo* signature estimation or the fitting of mutation counts to known signatures. Also, the app provides a module to explore public datasets from TCGA.

Start the app using either RStudio or a terminal:

```
# running signeRFlow function
signeRFlow()
```

The app will open on a new window or on a tab at your browser.

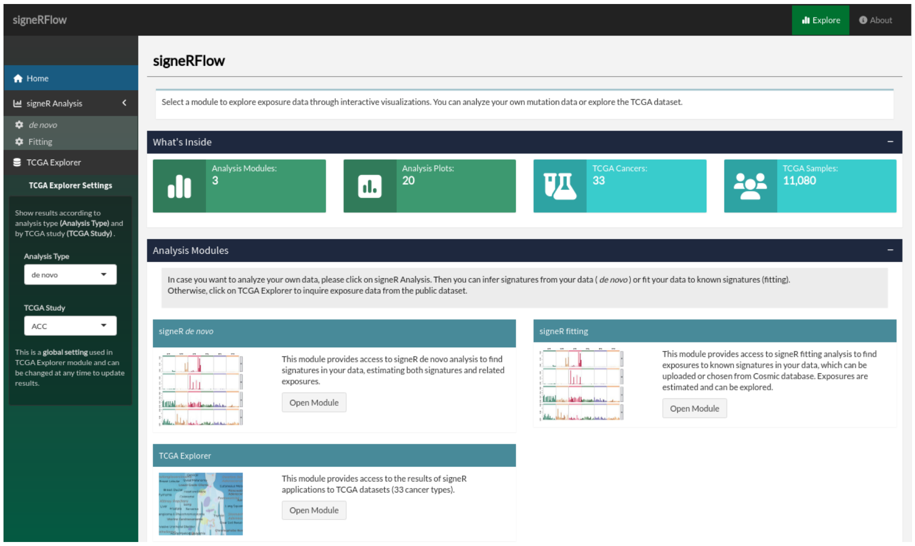


Figure 1: signeRFlow app overview.

### 2 Modules

signeRFlow functionalities are divided in three modules:

- signeR *de novo*: performs *de novo* analysis to extract signatures from your data, also estimating related exposures.
- signeR fitting: find exposures to known signatures in your data, which can be uploaded or chosen from Cosmic database. Exposures are estimated and can be explored.
- TCGA explorer: provides access to the results of signeR applications to 33 datasets from TCGA.

You can go through the modules independently by using the app sidebar. All modules give access to downstream analysis of exposure data.

#### 2.1 signeR *de novo* analysis

In this module, you can upload a SNV matrix with counts of mutations and execute the signeR *de novo* algorithm, which computes a Bayesian approach to the non-negative factorization (NMF) of the mutation counts in a matrix product of mutational signatures and exposures to mutational processes.

You can also provide a file with opportunities that are used as weights for the factorization. Further analysis parameters can be set, results can be visualized on different plots and found signatures can be compared to the ones in Cosmic database interactively.

##### 2.1.1 Load data

You can upload a VCF file or a SNV matrix file with your own samples to use in signeR *de novo* module. You can upload an opportunity file as well or use a already built genome opportunity. Also, you can upload a BED file to build an opportunity matrix.

#### VCF or SNV matrix

You can upload a VCF file or a SNV matrix file from your computer by clicking at the **Browse button**.

SNV matrix is a text file with a (tab-delimited) matrix of SNV counts found on analyzed genomes. It must contain one row for each genome sample and 97 columns, the first one with sample ids and, after that, one column for each mutation type. Mutations should be specified in the column names (headers), by both the base change and the trinucleotide context were it occurs (for example: C>A:ACA). The table below shows a example of the SNV matrix structure.

If you want to upload a VCF file, you must select the genome build used on your variant calling analysis to allow signeR to generate a SNV matrix of counts. Also, you can generate a SNV matrix from a VCF file using the method:

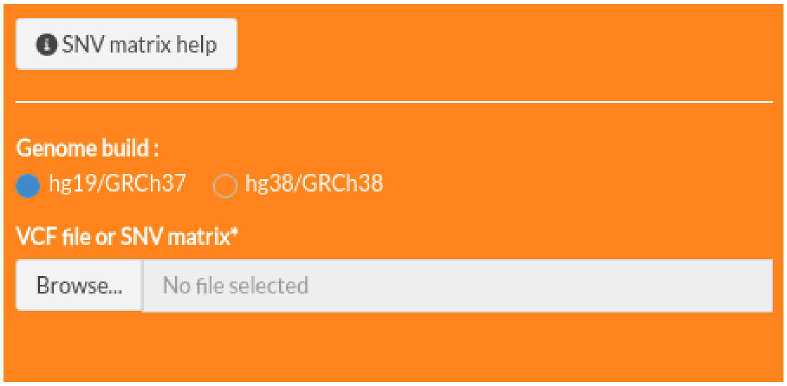


Figure 2: VCF or SNV matrix upload.

Table 1: SNV matrix example.

| | C>A:ACA | C>A:ACC | C>A:ACG | C>A:ACT | C>A:CCA | ... | T>G:TTT |
|---|---|---|---|---|---|---|---|
| PD3851a | 31 | 34 | 9 | 21 | 24 | ... | 21 |
| PD3904a | 110 | 91 | 9 | 87 | 108 | ... | 77 |
| ... | ... | ... | ... | ... | ... | ... | ... |
| PD3890a | 122 | 112 | 13 | 107 | 99 | ... | 50 |

`genCountMatrixFromVcf`

from signeR package. See the documentation for more details.

**Columns**:
The first column needs to contain the sample ID and other columns contain the 96 trinucleotide contexts.

**Rows**:
Each row contains the sample ID and the counts for each trinucleotide contexts.

#### Opportunity matrix

You can upload an Opportunity matrix file or a BED file from your computer by clicking at the **Browse button**. Also, you can use a already built genome opportunity for human reference genomes. This is an optional file.

Opportunity matrix is a tab-delimited text file with a matrix of counts of trinucleotide contexts found in studied genomes. It must structured as the SNV matrix, with mutations specified on the head line (for each SNV count, the Opportunity matrix shows the total number of genomic loci where the refereed mutation could have occurred). The table below shows a example of the opportunity matrix structure.

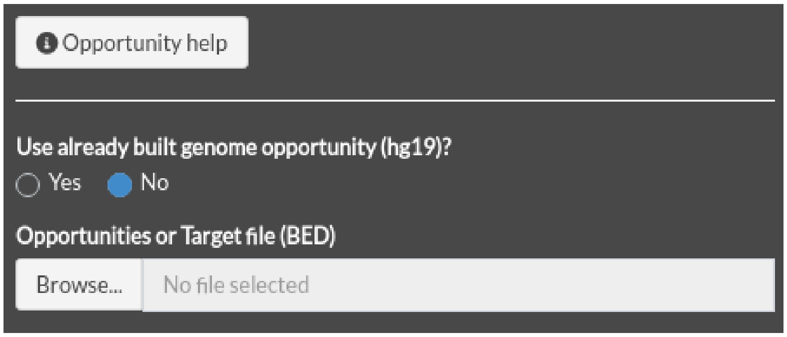


Figure 3: Opportunity matrix or BED upload.

Table 2: Opportunity matrix example

| | | | | | | | |
|---|---|---|---|---|---|---|---|
| 366199887 | 211452373 | 45626142 | 292410567 | 335391892 | 239339768 | ... | 50233875 |
| 202227618 | 116207171 | 25138239 | 161279580 | 184193767 | 131051208 | ... | 177385805 |
| 225505378 | 130255706 | 28152934 | 179996700 | 206678032 | 147634427 | ... | 199062504 |
| 425545790 | 245523433 | 53437284 | 339065644 | 389386002 | 278770926 | ... | 375075216 |
| 452332390 | 259934779 | 55862550 | 361010972 | 412168035 | 292805460 | ... | 396657807 |

If you want to upload a BED file, you must select the genome build used on your analysis to allow signeR to generate the opportunities for your regions file. Also, you can generate an opportunity matrix from the reference genome using the method:

`genOpportunityFromGenome`

from signeR package. See the documentation for more details.

**Columns**:
There is no header in this file and each column represents a trinucleotide context.

**Rows**:
Each row contains the count frequency of the trinucleotides in the whole analyzed region for each sample.

###### 2.1.2 *De novo* analysis parameters

There are some parameters that you can define before running the analysis by clicking at **Start *de novo* analysis** button:

Parameters:

- **Number of signatures**: define the minimal and maximal numbers of signatures you want that signeR estimates.

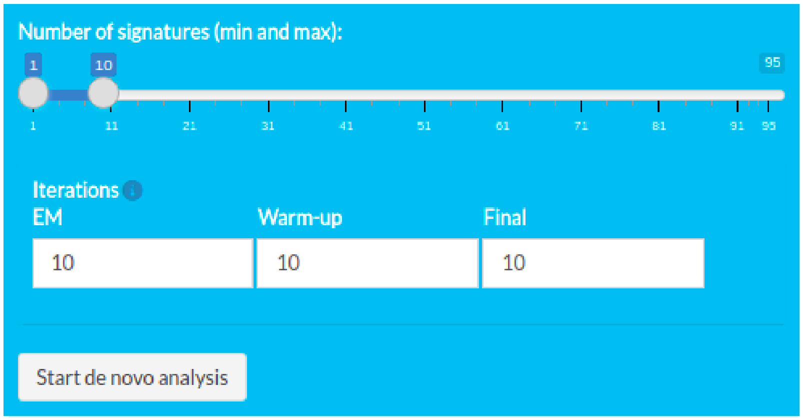


Figure 4: de novo analysis.

- **EM**: number of iterations performed to estimate the hiper-hiper parameters of signeR model. Ignored if previously computed values are used for those parameters (fast option).

- **Warm-up**: number of Gibbs sampler iterations performed in warming phase, before signeR assumes that the model have converged.

- **Final**: number of final Gibbs sampler iterations used to estimate signatures and exposures.

During the execution, a message will appear at the screen showing the progress. After the analysis is finished, you can download the results by clicking the button **Download Rdata** below the button **Start de novo analysis** and can iterate with all available plots in signeR package. For example, the convergence of the MCMC model used to estimate the signatures along with their exposures can be seen on Figure 5.

###### 2.1.3 Cosmic cosine

signeRFlow uses COSMIC v3.2 to calculate the cosine distance between found signatures and those present in COSMIC. A heatmap will be shown at the **COSMIC Comparison** section of *de novo* tab.

##### 2.2 signeR fitting

In this module, you can upload a VCF file or a SNV matrix with counts of mutations, the same as used on *de novo* module, and a previous signatures file with known signatures to execute the signeR fitting algorithm, witch computes a Bayesian approach to the fitting of mutation counts to known mutational signatures, thus estimating exposures to mutational processes.

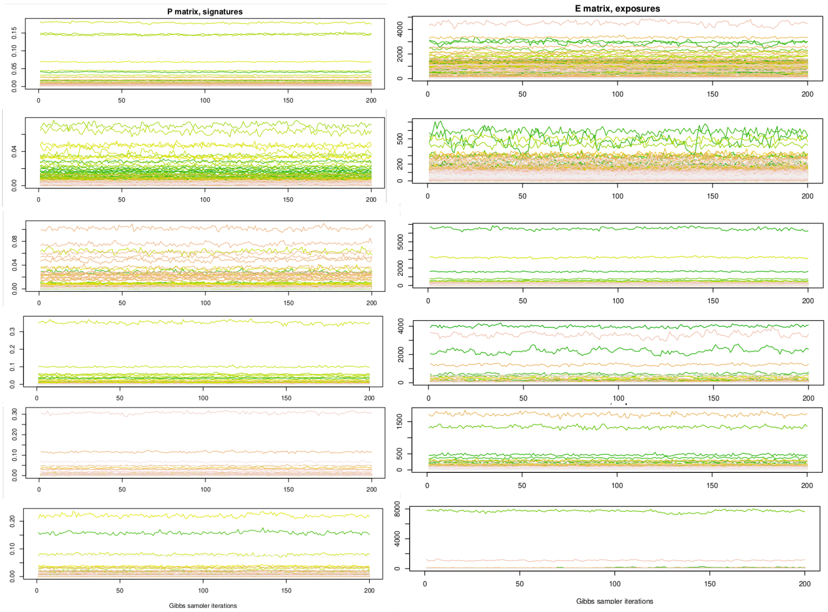


Figure 5: Estimated entries for both the signatures matrix and the exposure matrix, along iterations of the Gibbs sampler.

You can also provide a file with opportunities or use a already built genome opportunity that are used as weights for the factorization. Further analysis parameters can be set and estimated exposures can be visualized on different plots interactively.

##### 2.2.1 Load data

You can upload a VCF file or a SNV matrix file with your own samples to use in signeR fitting module and previous known signatures. You can upload an opportunity file as well. SNV or VCF (2.1.1) and opportunity (2.1.1) matrix are the same as used on *de novo* module.

#### Previous signatures matrix

You can upload a Previous signatures matrix file from your computer by clicking at the **Browse button**.

Previous signatures is a tab-delimited text file with a matrix of previously known signatures. It must contain one column for each signature and one row for each of the 96 SNV types (considering trinucleotide contexts). Mutation types should be contained on the first column, in the same form as the column names of the SNV (2.1.1) matrix. The table below shows a example of the previous signatures matrix structure.

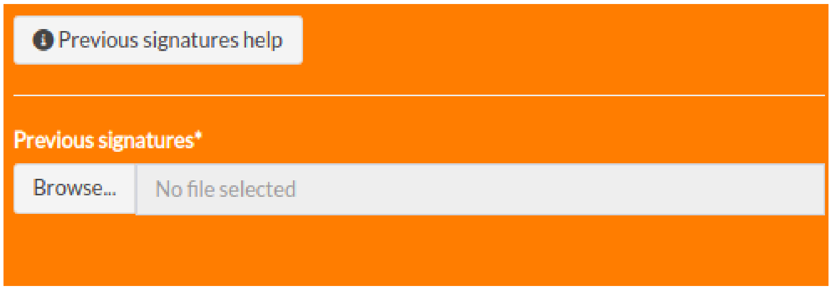


Figure 6: Previous known signatures matrix upload.

Table 3: Previous known signature example

| | Signature 2 | Signature 3 | Signature 5 | Signature 6 | ... | Signature 8 |
|---|---|---|---|---|---|---|
| C>A:ACA | 0.01110 | 0.00067 | 0.02218 | 0.01494 | ... | 0.03672 |
| C>A:ACC | 0.00915 | 0.00062 | 0.01788 | 0.00896 | ... | 0.03324 |
| C>A:ACG | 0.00150 | 0.00010 | 0.00213 | 0.00221 | ... | 0.00252 |
| ... | ... | ... | ... | ... | ... | ... |
| T>G:TTT | 0.00403 | 2.359E-05 | 0.0130 | 0.01337 | ... | 0.00722 |

**Columns**:
The first column needs to contain the trinucleotide contexts and other columns contain the known signatures.

**Rows**:
Each row contains the expected frequency of the given mutation in the appointed trinucleotide context.

###### 2.2.2 Fitting parameters

There are some parameters that you can define before running the analysis by clicking at **Start Fitting analysis** button:

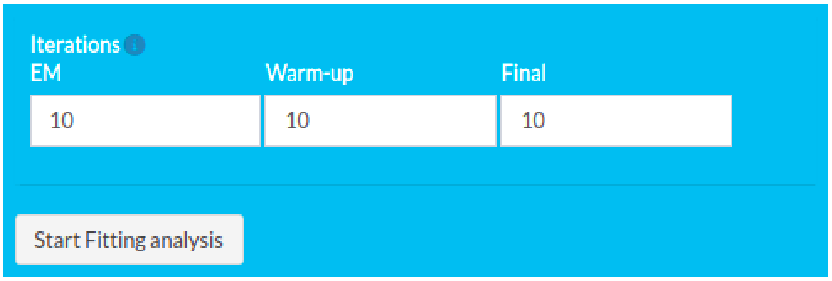


Figure 7: Fitting analysis

Parameters:

- **EM**: number of iterations performed to estimate the hiper-hiper parameters of signeR model. Ignored if previously computed values are used for those parameters (fast option).
- **Warm-up**: number of Gibbs sampler iterations performed in warming phase, before signeR assumes that the model have converged.
- **Final**: number of final Gibbs sampler iterations used to estimate signatures and exposures.

During the execution, a message will appear at the screen showing the progress.

#### 2.3 TCGA Explorer

Instead of uploading a private dataset, signeRFlow allows you to explore exposure data previously estimated for samples on TCGA public datasets. We previously applied the signeR algorithm to genome samples from 33 cancer types. Estimated mutational signatures and exposures were obtained for each cancer type. Also, known signatures from Cosmic database were fitted to TCGA mutation data, thus estimating related exposures on each cancer type.

You can select the cancer type of interest and the analysis type on the sidebar. Also, samples can be filtered according to available features in the metadata.

The first time you click in the button **TCGA Explorer** on the sidebar, signeRFlow will download all the necessary files (RData) according to cancer study and analysis type. The files are often small, but depends on the cancer study, this process can take a while. A message will show the download and rendering progress.

##### 2.3.1 Filter dataset

Using the data summary table with all clinical data features downloaded from TCGA, you can select a feature to filter the dataset. According to the feature class, different options to filter will be shown. If you filter a dataset using the data summary table, it will be used on the downstream analysis, such as clustering and covariate.

It is not mandatory to filter the dataset, you can use all the cases. The aim of this resource is to allow you to explore the dataset and select the cases you work with.

As an example, we selected the feature *ajcc_pathologic_stage* from ACC cancer type and *de novo* analysis:

and applied the filter on the dataset, selecting only groups Stage I and Stage II:

For each change on feature and filters, the available plots are updated according to the filtered samples.

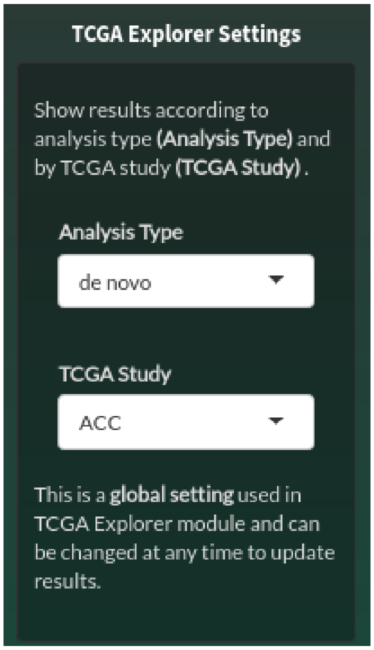


Figure 8: TCGA Explorer

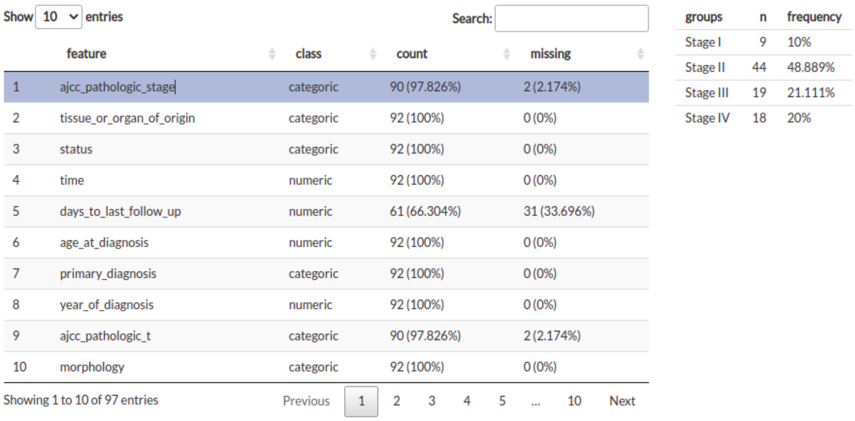

Show 10 entries Search:

| | feature | class | count | missing |
|---|---|---|---|---|
| 1 | ajcc_pathologic_stage | categoric | 90 (97.826%) | 2 (2.174%) |
| 2 | tissue_or_organ_of_origin | categoric | 92 (100%) | 0 (0%) |
| 3 | status | categoric | 92 (100%) | 0 (0%) |
| 4 | time | numeric | 92 (100%) | 0 (0%) |
| 5 | days_to_last_follow_up | numeric | 61 (66.304%) | 31 (33.696%) |
| 6 | age_at_diagnosis | numeric | 92 (100%) | 0 (0%) |
| 7 | primary_diagnosis | categoric | 92 (100%) | 0 (0%) |
| 8 | year_of_diagnosis | numeric | 92 (100%) | 0 (0%) |
| 9 | ajcc_pathologic_t | categoric | 90 (97.826%) | 2 (2.174%) |
| 10 | morphology | categoric | 92 (100%) | 0 (0%) |

Showing 1 to 10 of 97 entries Previous 1 2 3 4 5 ... 10 Next

| groups | n | frequency |
|---|---|---|
| Stage I | 9 | 10% |
| Stage II | 44 | 48.889% |
| Stage III | 19 | 21.111% |
| Stage IV | 18 | 20% |

Figure 9: TCGA Data summary

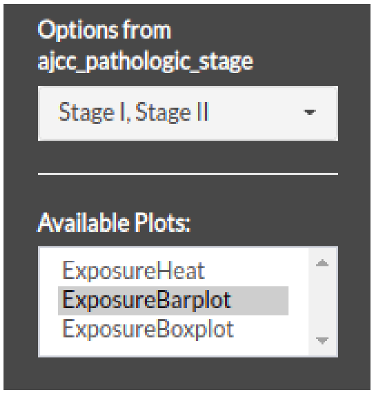


Figure 10: TCGA filter dataset

After, you can download the results by clicking the button **Download Rdata** below the button **Start Fitting analysis** and iterate with all available plots in signeR package.

### 3 Downstream analysis

Available in all modules, you can perform downstream analysis using *de novo* or fitting results with your own data, or in the TCGA Explorer module. Available analysis options, conditioned by the provided clinical data, will be found on the top tabs *Clustering* and *Covariate*. On TCGA Explorer module it is not needed to upload a clinical dataset, since available data is embedded in the software. In this last case, in the top of **Covariate** tab you will see, as a reminder, information about the dataset and used filters.

There are two main downstream analysis:

- **Clustering**
  - *Hierarchical Clustering*: signeRFlow generates a dendogram for each generated sample of the exposure matrix. Consensus results, i.e. branches that are recurrently found, are reported. Different distance metrics and clustering algorithms are available to be selected.
  - *Fuzzy Clustering*: signeRFlow can apply the Fuzzy C-Means Clustering on each generated sample of the exposure matrix. Pertinence levels of samples to clusters are averaged over different runs of the algorithm. Means are considered as the final pertinence levels and are shown in a heatmap.
- **Covariate**

– *Categorical feature*: differences in exposures among groups can be analyzed and if some of the samples are unlabeled they can be labeled based on the similarity of their exposure profiles to those of labeled samples.

– *Continuous feature*: its correlation to estimated exposures can be evaluated.

– *Survival feature*: survival data can also be analyzed and the relation of signatures to survival can be accessed.

##### 3.0.1 Clustering

###### Hierarchical Clustering

By using the Hierarchical clustering section, you can select different dist and hclust methods:

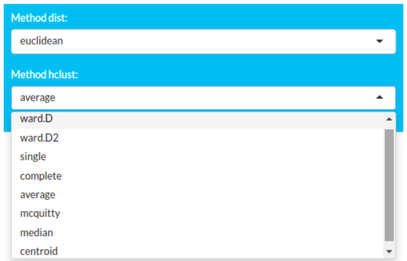


Figure 11: hclust methods

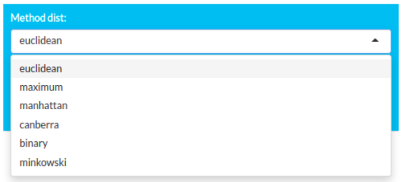


Figure 12: hclust distances

When you select a new *dist* or *hclust* method, a dendogram plot is updated.

###### Fuzzy Clustering

By using the Fuzzy clustering section, you can set the number of groups or let the algorithm to estimate (Set groups to 1) and click at the **Run fuzzy** to start the analysis:

During the execution, a message will appear at the screen showing the progress[1]. The output of Fuzzy clustering is shown as a heatmap plot.

##### 3.0.2 Covariate

To perform a Covariate analysis on signeRFlow, you must upload a clinical data, a tab-delimited file with samples in rows and features in columns. You can upload a file by clicking in the **Browse...** button:

Clinical data is a tab-delimited text file with a matrix of available metadata (clinical and/or survival) for each sample. It must have a first column of sample ids, named as **"SampleID"**, whose entries match the row names of the **SNV**

[1]Warning: Fuzzy clustering can be a long process and demands high computer resources.

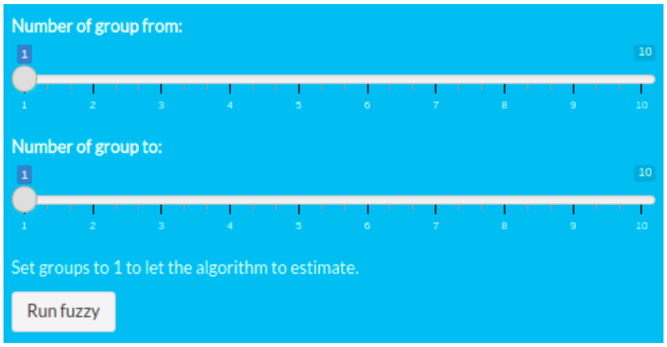


Figure 13: Fuzzy clustering

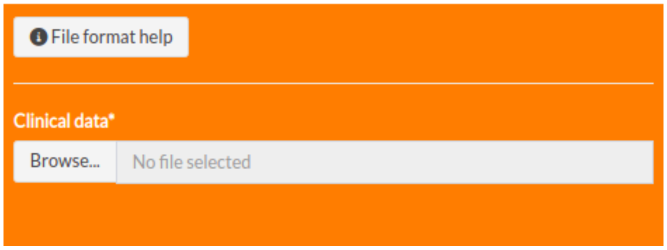


Figure 14: clinical data upload

**matrix**(2.1.1). The number and title of the remaining columns are optional, however if **survival** data is included it must be organized in a column named *time* (in months) and another named *status* (which contains 1 for death events and 0 for censored samples). The table below shows a example of the clinical data matrix structure.

Table 4: Clinical data matrix example

| SampleID | gender | ajcc_pathologic_stage | ethnicity | race | status | time |
|---|---|---|---|---|---|---|
| PD3851a | male | Stage I | not hispanic or latino | white | 0 | 236 |
| PD3890a | male | Stage II | not hispanic or latino | black or african american | 1 | 199 |
| PD3904a | female | Stage II | NA | NA | 0 | 745 |
| PD3905a | female | Stage IV | NA | white | 1 | 299 |
| PD3945a | male | Stage IV | not hispanic or latino | asian | 0 | 799 |

**Columns**:

The first column must contains the sample ID. Other columns may contain sample groupings or other features that you would like to co-analyze with exposure data.

**Rows**:

Each row contains clinical information for one sample: its ID and all other data of interest.

After the upload, a description table summarizes the data with all the features in rows, and the class, counts and missing for each feature. By selecting a feature (row) at the table, a small panel is shown next to the table summarizing the values, categorical or continuous, for the selected feature:

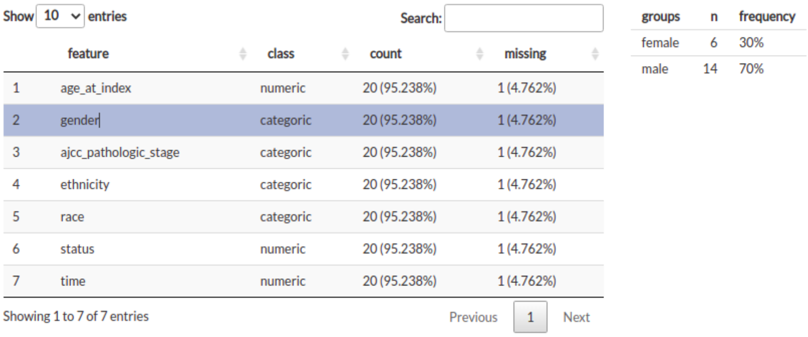

Show 10 entries Search:

| | feature | class | count | missing |
|---|---|---|---|---|
| 1 | age_at_index | numeric | 20 (95.238%) | 1 (4.762%) |
| 2 | gender | categoric | 20 (95.238%) | 1 (4.762%) |
| 3 | ajcc_pathologic_stage | categoric | 20 (95.238%) | 1 (4.762%) |
| 4 | ethnicity | categoric | 20 (95.238%) | 1 (4.762%) |
| 5 | race | categoric | 20 (95.238%) | 1 (4.762%) |
| 6 | status | numeric | 20 (95.238%) | 1 (4.762%) |
| 7 | time | numeric | 20 (95.238%) | 1 (4.762%) |

Showing 1 to 7 of 7 entries Previous 1 Next

| groups | n | frequency |
|---|---|---|
| female | 6 | 30% |
| male | 14 | 70% |

Figure 15: Clinical data summary

According to the class of the feature, a set of analysis are available in the **Plots** section:

##### *Categorical features*

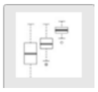
**Differential Exposure Analysis**: highlight signatures that are differentially active among groups of samples.

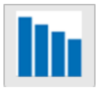
**Sample Classification**: classify samples based on their exposures to mutational processes.

##### *Numeric features*

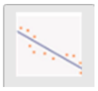
**Correlation Analysis**: evaluates the correlation of exposure levels for each mutational signature to provided feature.

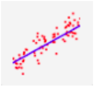 **Linear Regression**: Builds a linear model of provided feature based on exposure data. Evaluates the relevance of each signature exposures in final model.

***Survival feature***

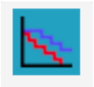 **Survival Analysis**: evaluate the effect of exposure levels to each signature on survival.

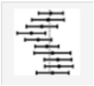 **Cox Regression**: uses a Cox Proportional Hazards Model to evaluate the combined effect on survival of exposure levels to different signatures.

### 4 Case study

The utility of signeRFlow app was demonstrated by its application on the data obtained from the stomach adenocarcinoma cohort available in TCGA (STAD, N=439). The mutational spectra of those samples were fitted to known mutational signatures found on COSMIC and previously described in this type of tumors (COSMIC v2; SBS1, SBS2, SBS5, SBS13, SBS15, SBS17, SBS18, SBS20, SBS21, SBS26 and SBS28). Part of found results were shown in the main paper and part are described below.

#### 4.1 Correlation analysis

signeR 2.0 can evaluate the correlation of continuous sample features and exposures to mutational signatures (ExposureCorrelation, Figure 16). For the STAD samples, the age at diagnosis were picked to study as example. COSMIC signatures SBS1, 5, 15, 20 and 26 showed significant Spearman's rank correlation with age, what can be explained by their etiology: SBS1 is related to endogenous mutational process and is expected to correlate with age; SBS15, 20 and 26 are associated with defective DNA mismatch repair.

#### 4.2 Linear regression

The software can also build generalized linear models of studied features based on their exposures to mutational processes, exporting the relevance of each signature in the model (ExposureGLM, Figure 17). The age at diagnosis were also used as example here, however linear models based on the exposures were not able to predict patients age satisfactorily.

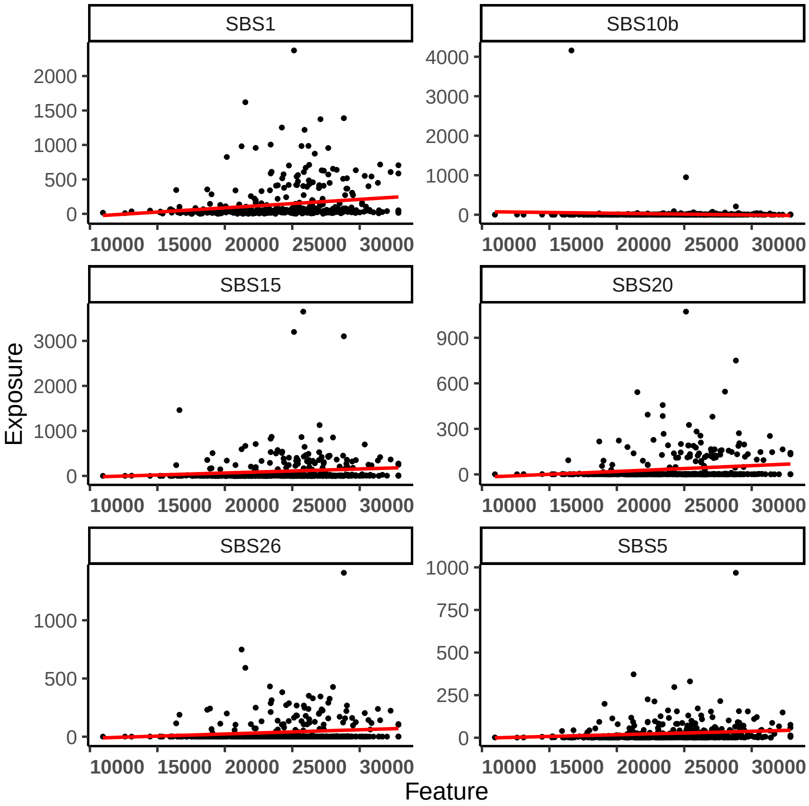


Figure 16: Scatter plots of exposures vs age at diagnosis.

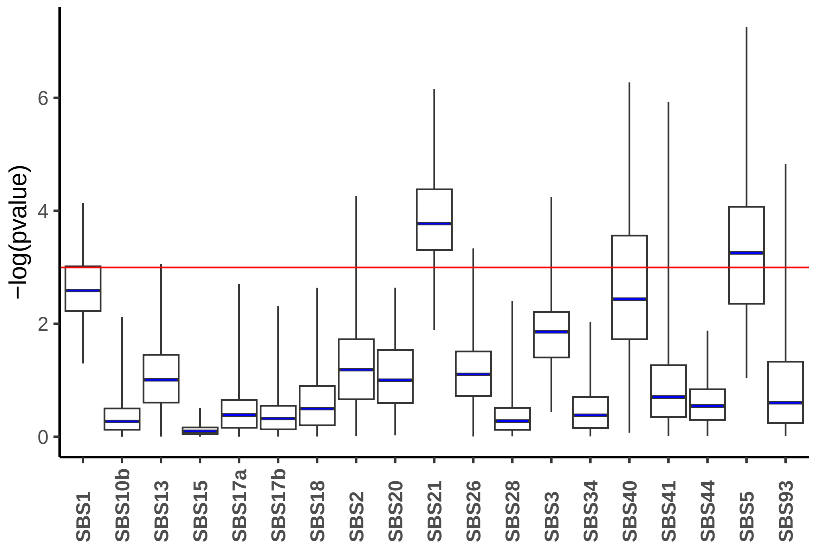


Figure 17: P-values boxplots showing the relevance of each signature's exposures to the modeling of age at diagnosis.

#### 4.3 Survival analysis

Survival data are extremely valuable in cancer studies. If those are available, exposure effects on overall survival can be analyzed for each signature. Log-rank tests and Cox proportional models are implemented and ready to be applied on exposure data from each signature. (ExposureSurv, Figure 18). Using the signeRFlow interface to select data before running the tests, we restricted the survival analysis of TCGA STAD samples to the ones classified as MSI-High. Among those, COSMIC signatures SBS1, 5, 15, 20, 21 and 26 were found as significantly related to better overal suvival. That founds are in accordance both to literature and to the previous result (age correlation test): apparently, signatures SBS1, 5, 15, 20, and 26, age at diagnosis and overall survival are closely related.

#### 4.4 Cox Regression

The combined effect of signature exposures on overall survival can also be assessed by a multidimensional Cox Proportional Hazards model. As can be seen on Figure 19, when considered all together no signature shows significant impact on survival.

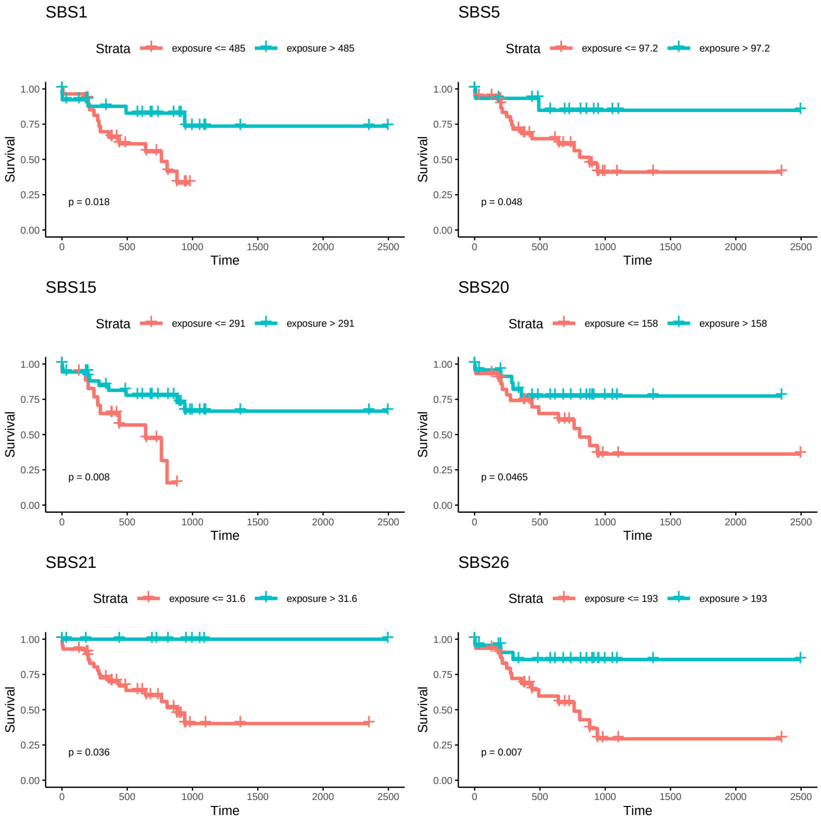


Figure 18: Kaplan-Meier curves showing survival differences in sample groups stratified by signatures exposures.

| | HR(CI) | P.value |
|---|---|---|
| SBS1 | 0.503 (0.135−1.988) | 0.315 |
| SBS2 | 1.241 (0.852−1.83) | 0.226 |
| SBS3 | 1.008 (0.782−1.289) | 0.443 |
| SBS5 | 0.954 (0.758−1.229) | 0.449 |
| SBS10b | 1.034 (0.771−1.392) | 0.355 |
| SBS13 | 0.932 (0.633−1.378) | 0.348 |
| SBS15 | 0.976 (0.368−2.401) | 0.581 |
| SBS17a | 0.929 (0.676−1.305) | 0.418 |
| SBS17b | 0.835 (0.588−1.167) | 0.23 |
| SBS18 | 0.899 (0.68−1.182) | 0.369 |
| SBS20 | 0.921 (0.499−1.765) | 0.411 |
| SBS21 | 0.865 (0.667−1.117) | 0.234 |
| SBS26 | 0.808 (0.477−1.339) | 0.345 |
| SBS28 | 0.983 (0.662−1.449) | 0.33 |
| SBS34 | 0.936 (0.637−1.386) | 0.348 |
| SBS40 | 0.912 (0.65−1.297) | 0.352 |
| SBS41 | 0.945 (0.644−1.408) | 0.387 |
| SBS44 | 0.952 (0.762−1.208) | 0.359 |
| SBS93 | 1.001 (0.664−1.494) | 0.388 |

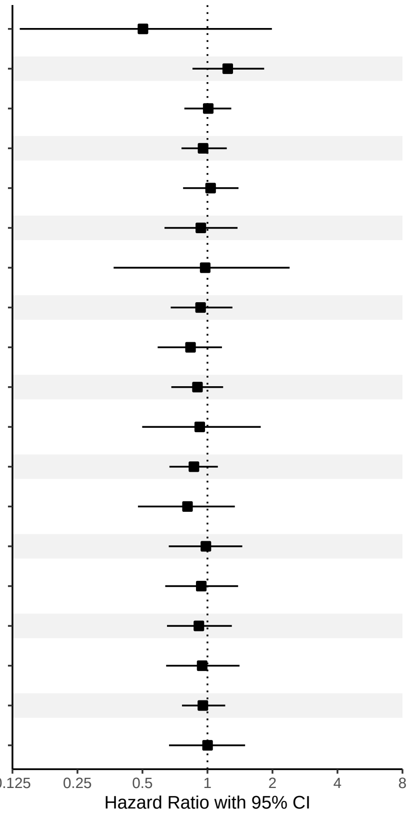


Figure 19: Forestplot showing the effect on survival of exposure levels for each signature.
